# Hasty Reduction of COVID-19 Lockdown Measures Leads to the Second Wave of Infection

**DOI:** 10.1101/2020.05.23.20111526

**Authors:** Yara Hazem, Suchitra Natarajan, Essam R. Berikaa

## Abstract

The outbreak of COVID-19 has an undeniable global impact, both socially and economically. March 11th, 2020, COVID-19 was declared as a pandemic worldwide. Many governments, worldwide, have imposed strict lockdown measures to minimize the spread of COVID-19. However, these measures cannot last forever; therefore, many countries are already considering relaxing the lockdown measures. This study, quantitatively, investigated the impact of this relaxation in the United States, Germany, the United Kingdom, Italy, Spain, and Canada. A modified version of the SIR model is used to model the reduction in lockdown based on the already available data. The results showed an inevitable second wave of COVID-19 infection following loosening the current measures. The study tries to reveal the predicted number of infected cases for different reopening dates. Additionally, the predicted number of infected cases for different reopening dates is reported.

## Introduction

The COVID-19 (SARS-CoV-2 virus) pandemic is a global challenge that requires accurate forecasting and prediction methods to address its socio-economic implications [1]. Globally, governments have imposed firm lockdown conditions and mandatory quarantine measures to reduce the spreading of the virus [2; 3]. These measures have successfully reduced the infection rate in many countries; however, they have significant negative socio-economic impacts [4]. Hence, it is important to understand the consequences of easing lockdown measures and refreshing the economy. Currently (up to 15th of May 2020), the countries can be classified into 3 categories: (A) countries that have successfully conquered the COVID-19 pandemic such as China, South Korea, and Australia, (B) countries on their way of beating COVID-19 and have already reached their peak infection period such as the United States, Italy, and Spain, and (C) countries that have not yet reached their peak infection period such as Russia, India, and Brazil [5]. Countries from both (A) and (B) categories have started working on outlining their plans to reopen and save their economy [6].

This paper aims to study the implications of loosening the lockdown conditions in the United States, Germany, the United Kingdom, Italy, Spain, and Canada. All the chosen countries have already passed their peak infection period, and their governments have announced their intentions of reducing the lockdown measures and reopening some nonessential services within the next months. A modified version of the Susceptible, Infectious, and Recovered (SIR) model is used to accurately simulate the pandemic [7]. For the study in hand, this model is used to forecast the infection rate if the lockdown measures are reduced by 25% on the 1st of June 2020 or the 1st of July 2020; hence, the impact of delaying this step is also investigated. This study gives a quantitative point of view for the risks of hasty reduction in lockdown measures and predicts the evolution of the number of infected cases until the end of 2020 following the assumed conditions.

## Methodology

Several mathematical and physical models describe the evolution of an epidemic; these models differ in capabilities and complexity [7; 8; 9; 10; 11]. In this work, the SIR model is used due to its flexibility and simplicity, which make it the commonly adopted model in the literature [12]. However, the SIR basic formulation is not capable of modeling the variations in the lockdown measures and its impact on the infection rate. Recent studies confirmed the accurate modeling of the COVID-19 epidemic with modified versions of the SIR model in all infection hot spots [13; 14; 15]. The basic formalism of the SIR model is depicted in the following coupled set of differential equations:

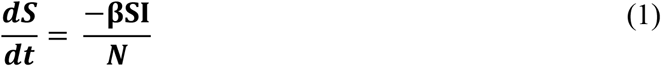

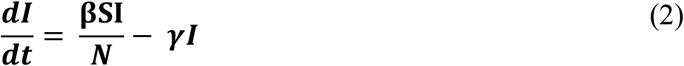

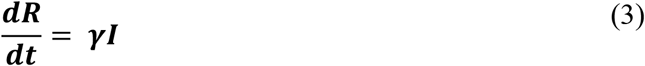

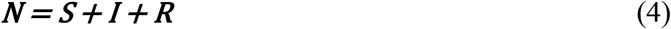

Where S indicates the time-dependent susceptible population, I is the time-dependent number of infected cases, R is the time-dependent number of recovered cases, N is the total population size, β resembles the probability of infection transmission from an infected case to a susceptible case, and *γ* resembles the rate at which infected cases recover. The initial conditions of the model are the whole population being susceptible to infection (S(t_0_)), the number of reported infected cases on the starting date of the outbreak (I(t_0_)), and zero recovered cases (R(t_0_)).

COVID-19 data is obtained from Johns Hopkins Center for Systems Science and Engineering (CSSE) data respiratory [16; 17], the data used in this study spans the interval between 20th of January 2020 until 15th of May 2020. The proposed algorithm to investigate the impact of reducing lockdown measures on infection rate is shown in Fig. 1. The acquired data is analyzed to extract the infection rate per country. The outbreak date and turning point of the infection rate are extracted in order to divide the data into 2 intervals: the first interval is characterized by a high infection rate and corresponds to low lockdown measures, and the second interval is characterized by lower infection rate and reflects the impact of the governmental lockdown measures. The high infection rate at the first interval can be interpreted by the lack of testing kits, social awareness, and governmental quarantine measures, which leads to a high β value.

**Fig 1.**
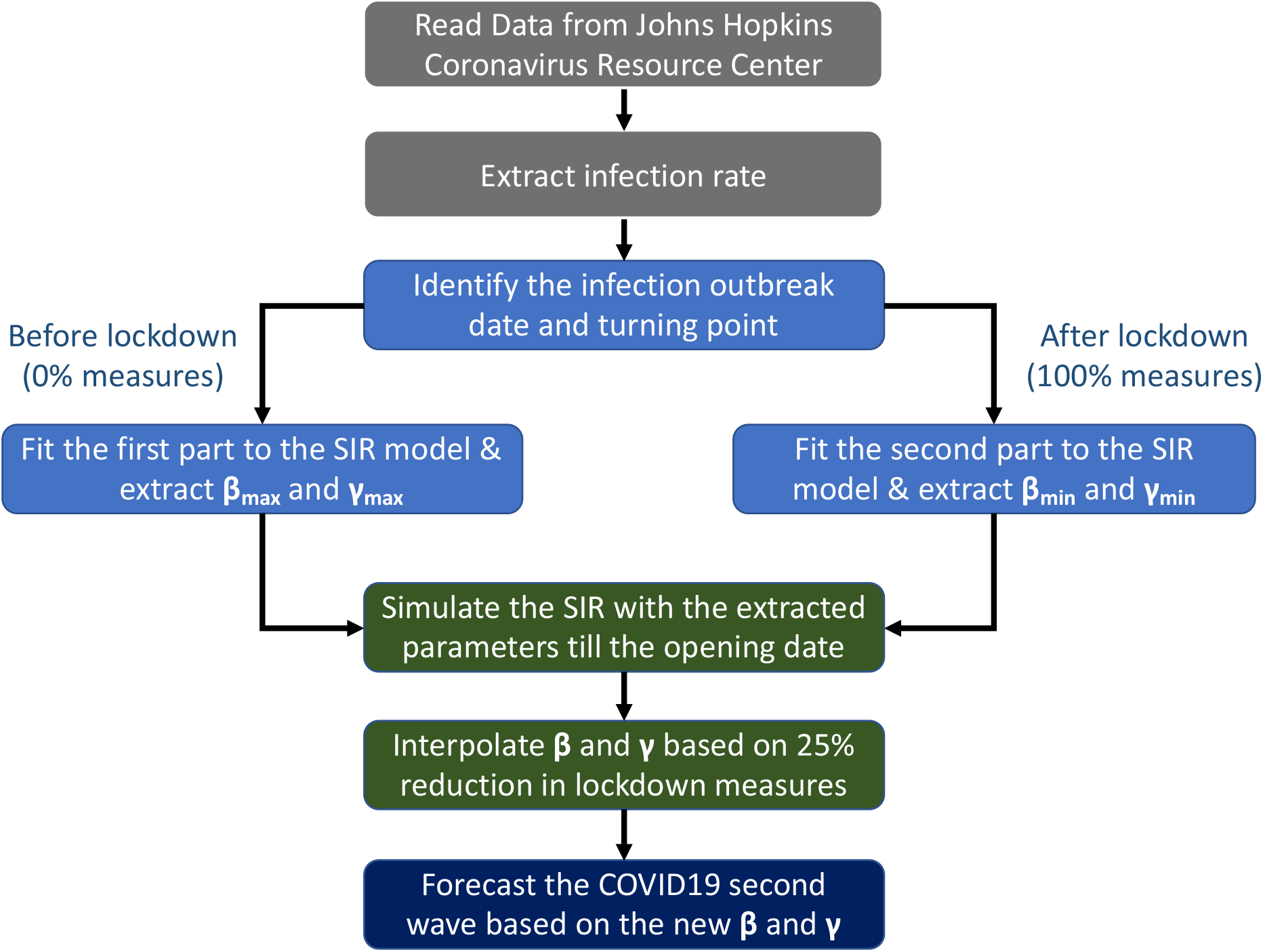
Flowchart of the proposed algorithm used in predicting the impact of reducing lockdown measures

Additionally, the healthcare system during this period operates at full power, which is reflected in the slightly higher *γ* value. After the turning point and experiencing peak infection rate, the quarantine measures are strictly applied, and the testing kits have been developed with higher reliability and shorter waiting times [18]; thus, β decreases. During the peak of the pandemic, the overwhelming load on the health care system and medical staff leads to the crash of some hospitals either due to the spread of the virus and the infection of the doctors and nurses, or the lack of intensive care units for everyone. This load reduces the overall healthcare system capability and interprets the slight decrease in *γ* value [19]. The concept of time-dependent β and *γ* has been proposed previously to overcome the limitations of the SIR model and simulate the impact of the quarantine measures [14; 20]. Table 1 shows the parameters extracted from the SIR model after fitting it to the acquired COVID-19 data for the considered countries in the study.

**Table 1.** The parameters used in calculations and forecasting number of new infected cases.

|  | United States | Germany | United Kingdom | Italy | Spain | Canada |
| --- | --- | --- | --- | --- | --- | --- |
| Population (Millions) | 328 | 83 | 66.65 | 60.36 | 46.94 | 37.59 |
| Outbreak date (d-m-yr) | 7-3-20 | 1-3-20 | 4-3-20 | 23-3-20 | 6-3-20 | 13-3-20 |
| Turning point (d-m-yr) | 7-4-20 | 28-4-20 | 14-4-20 | 22-4-20 | 27-4-20 | 19-4-20 |
| $\beta_{\max}$ | 14.5 | 19 | 13 | 9 | 10 | 8.5 |
| $\gamma_{\max}$ | 14.3 | 18.81 | 12.85 | 8.829 | 9.82 | 8.42 |
| $\beta_{\min}$ | 3.5 | 10.5 | 5.5 | 5.2 | 4.5 | 6 |
| $\gamma_{\min}$ | 3.45 | 10.39 | 5.42 | 5.17 | 4.48 | 5.97 |
| $\beta$ @ 25% reduction in measures | 4.36 | 11.26 | 6.21 | 5.68 | 5.1 | 6.37 |
| $\gamma$ @ 20% recovery of health care systems | 4.09 | 10.96 | 5.95 | 5.53 | 4.92 | 6.25 |
| Integrated number of newly infected cases (Thousands): |  |  |  |  |  |  |
| Reopen on 1st of June | 5800 | 94 | 380 | 163 | 227 | 80 |
| Reopen on 1st of July | 5500 | 91 | 345 | 133 | 217 | 55 |

Quantifying the governmental measures to compact the COVID-19 pandemic is difficult; we assumed that the first interval and the second interval correspond to 0% and 100% measures, respectively. These are just estimates; however, they can be used to predict the impact of loosening the lockdown measures on β and γ that in turn affects the infection rate. In this study, we considered loosening the lockdown measures by 25% compared to the 100% measures during the second interval. The infection rate has an exponential dependence on β and *γ* [20]; thus, an exponential dependence on the lockdown measures is assumed, and the parameters used for prediction are obtained according to:

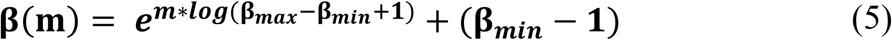

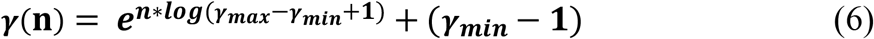

Where m corresponds to the reduction percentage of the lockdown measures, n corresponds to the recovery percentage of the healthcare system, β*_max_* and *γ_max_* are the parameters of the first interval, and β*_min_* and *γ_min_* correspond to the parameters of the second interval. The calculated values are reported in Table 1. Further, the interpolated β and *γ* are used in forecasting the infection rate if the lockdown measures are reduced on the 1st of June 2020 or the 1st of July 2020. The increase in β models the reduction in lockdown measures, and the slight increase in *γ* models the partial recovery of the healthcare system. The continuity of the infection rate is imposed by the continuity of the susceptible and recovered populations.

## Results and Discussion

Based on the described modeling scheme, the increase in infection rate due to loosening of lockdown measures is calculated until the end of 2020, as shown in Fig. 2. Countries react differently to the proposed scheme as revealed by Fig. 2 and the values of the predicted number of infected cases tabulated in Table 1. Qualitatively, loosening the measures shall result in a significant increase in the number of infected cases; however, this increase is dependent on the status of the country at the date of reopening. The United States and the United Kingdom are more affected by reopening as they have not fully conquered COVID-19 yet; hence, the hasty reduction of quarantine measures might lead to even higher infection rates that has happened before during the Spanish flu [21; 22]. Thus, there is a correlation between the turning point date, reopening date, and second-wave peak infection rate. Quantitively, this study reveals that the second wave might lead to 5.8 million newly infected cases in the United States, 94 thousand in Germany, 380 thousand in the United Kingdom, 163 thousand in Italy, 227 thousand in Spain, and 80 thousand in Canada. These values are based on the SIR predictions according to the aforementioned algorithm.

**Fig 2.**
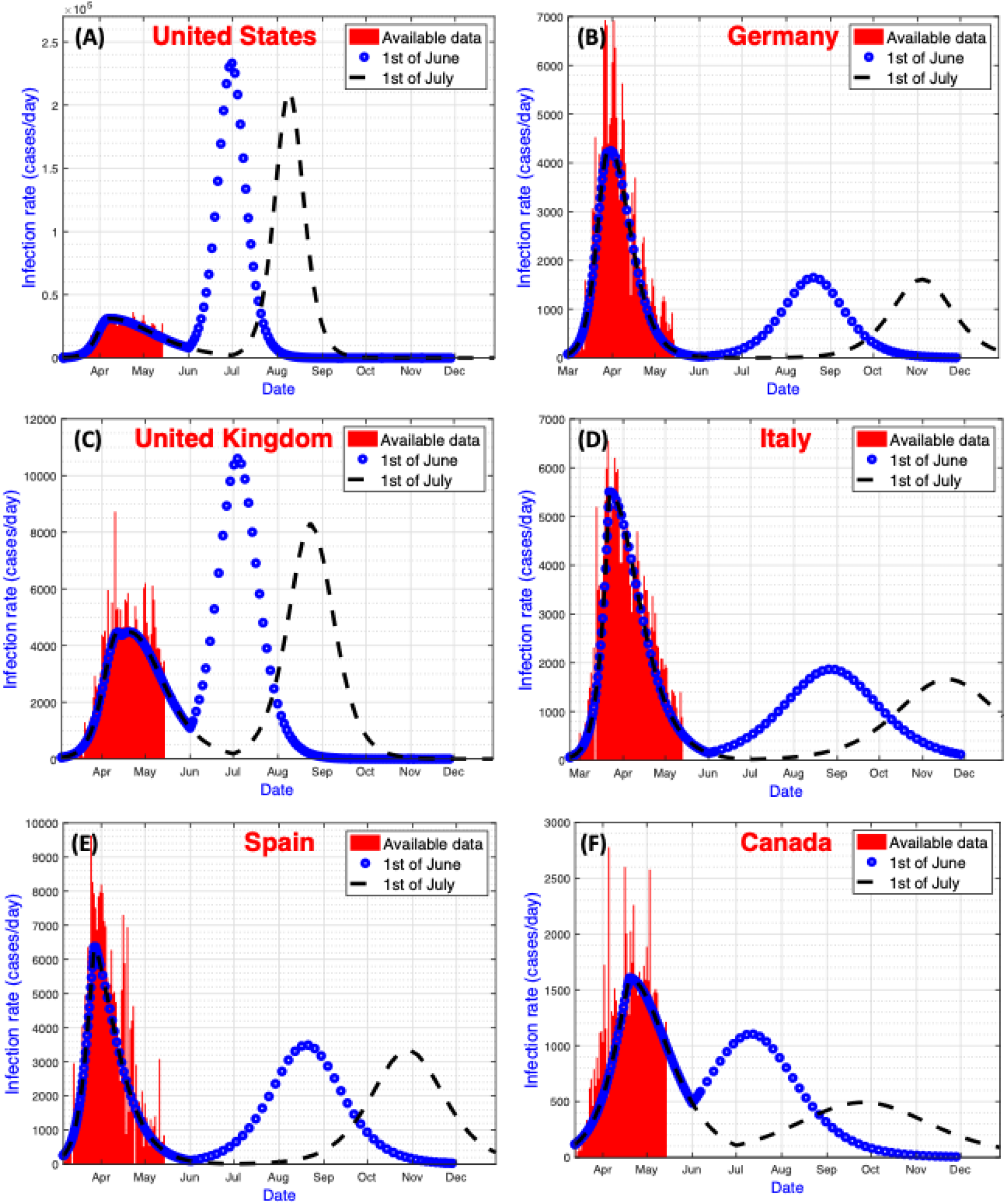
The evolution of the infection rate and the fitted SIR model. Red indicates the available COVID-19 data, and blue dotted line and black dashed line indicate the predicted infection rate if lockdown measures are reduced by 25% on the 1st of June 2020 and 1st of July 2020, respectively. Different plots correspond to different countries, according to (A) United States, (B) Germany, (C) United Kingdom, (D)Italy, (E) Spain, and (F) Canada

Based on the results presented in Table 1, it is recommended to consider loosening the measures after at least 3 months from the turning point date. Additionally, delaying the reopening from June 1st to July 1st can reduce the overall number of newly infected cases by 5% as in the United States and up to 45% as in Canada, which ensures that the capacity of the recovering healthcare system can meet the number of newly infected cases. According to Washington University COVID-19 model, the number of ICU beds and invasive ventilators needed are projected to continue to decrease until the end of August [23]. This decrease proves that the health care system will be able to partially recover before the second peak of the COVID-19 hits the countries.

Despite our best effort in analyzing and optimizing the data collected, there are a few limitations to be considered. Our model assumes that the partially recovered healthcare system is not deteriorated by the second wave; hence *γ* is fixed. In addition, the SIR model assumes that the recovered cases gain immunity against the disease, which is not the case for the COVID-19 pandemic [24]. Not forgetting that, the development of testing kits, treatment techniques, and a reliable vaccine can have a considerable positive impact on the infection rates, which is not considered in our analysis.

## Conclusion

In conclusion, this study offers a quantifiable prediction of how reducing the lockdown measures shall lead to the second wave of COVID-19 in the United States, Germany, the United Kingdom, Italy, Spain, and Canada. Further, delaying the reduction of lockdown measures show a reasonable reduction in the number of predicted infected cases. Eventually, this study highlights the risks of hasty reduction of lockdown measures in countries in the middle of their battle with COVID-19. This study aims mainly to ring alarm bells for the risks accompanied by the rash loosening of the quarantine measures. Moreover, this paper introduces a comprehensive worst-case view of shifting into the heard immunity paradigm.

Our calculations ignored any external measures to combat the COVID-19 second wave. For further studies, it is recommended to include the possibility of governmental interventions that might lead to a different scenario with peak predictions. In addition, it is very recommended to start evaluating the pandemic second wave, applying a time-varying β and *γ* factors.

## Data Availability

The study is based on the Modelling of the COVID-19 Pandemic, the data are already available from different sources as WHO.

https://github.com/CSSEGISandData/COVID-19

